# E-cigarette use frequency by smoking status among youth in the United States, 2014-2019

**DOI:** 10.1101/2020.05.07.20094185

**Authors:** Jamie Tam

## Abstract

**Aims:** To report annual 2014-2019 youth estimates of past 30-day e-cigarette use frequency by smoking status in the United States (US).

**Design:** Weighted prevalence estimates of student’s e-cigarette use using the 2014-2019 National Youth Tobacco Surveys (NYTS). For each year, t-tests for significance were used to compare estimates with those from the preceding year; t-tests were not performed on data for 2019 due to the change in survey format from paper to electronic.

**Setting:** The NYTS is an annual school-based cross-sectional survey of US middle school (MS) and high school (HS) students.

**Participants:** 117,472 students.

**Measurements:** Self-report of past 30 day e-cigarette use based on students’ smoking status. Smoking status is assessed by asking if students have ever tried smoking, “even one or two puffs”, with never smokers responding “no”. Former smokers respond “yes” but have not smoked at all in the past 30 days. Current smokers used cigarettes at least once in the past 30 days. Frequent e-cigarette use is defined as use on ≥20 days in the past month.

**Findings:** Past 30-day and frequent e-cigarette use increased among never, former, and current smoker youth from 2014-2019. In 2019, a greater proportion of current smokers used e-cigarettes frequently (HS = 46.1%, 95% CI: 39.1, 53.2; MS = 27.4%, 95% CI: 21.1, 33.6) compared to former smokers (HS = 23.2%, 95% CI: 18.1, 28.2; MS = 10.9%, 95% CI: 6.1, 15.7) and never smokers (HS = 3.7%, 95% CI: 3.0, 4.3; MS = 0.7%, 95% CI: 0.4, 0.9). From 2018 to 2019, the total number of youth using e-cigarettes frequently who were never smokers (2018: 180,000; 2019: 490,000) or former smokers (2018: 260,000; 2019: 640,000) surpassed that of current smokers (2018: 420,000; 2019: 460,000).

**Conclusions:** The proportion and number of never smoker youth using e-cigarettes frequently increased greatly since 2014.

## Introduction

Surveys show unprecedented increases in e-cigarette use among United States (US) youth since 2017.(1-4) High school students’ e-cigarette use increased in 2018, with 20.8% of high school students reporting use of an e-cigarette within the past month.(2) In 2019, that increased to 27.5%.(4) Yet patterns of e-cigarette use among youth are known to differ dramatically by smoking status, with much higher rates of use among current smokers compared to never smokers.(5-7) E-cigarette use is more common among youth who have already used other tobacco products.(8) For young people who smoke cigarettes and use e-cigarettes, their cigarette use presents their largest health risk. However, e-cigarettes present new health risks for young people who have never smoked or used tobacco products. Data from 2018 National Youth Tobacco Survey (NYTS) show that use of e-cigarettes rose among youth who were naïve to other tobacco products.(9) However the 2018 NYTS data likely underestimated e-cigarette use due to the exclusion of the popular brand JUUL from the list of examples for e-cigarettes— only in 2019 did the survey begin to explicitly mention ‘JUUL’ as a type of e-cigarette.(4)

In the US, public health leaders are especially concerned that never smoker youth might become addicted to nicotine through e-cigarettes, and that flavors could be the “on-ramp” to addiction.(10) In response to rising e-cigarette use among youth and an outbreak of e-cigarette lung injuries, several states have enacted emergency rules to temporarily restrict sales of flavored e-cigarettes, and the Food and Drug Administration has removed certain flavored cartridge-based e-cigarettes, such as JUUL.(11,12)

For never smoking youth who use e-cigarettes, the extent to which they are at risk also depends on their frequency of exposure to nicotine. Research from 2014-2015 showed that high e-cigarette use frequency was concentrated predominately among youth who already use combustible tobacco products, with very low levels of frequent e-cigarette use among never smokers.(7, 8,13) Other data from the 2014 Monitoring the Future survey also showed that among non-smoking high school students who use e-cigarettes, most used them on 1-2 of the past 30 days.^3^ Frequent use of e-cigarettes is associated with nicotine dependence among youth,(14) however never smokers who are using e-cigarette infrequently may not be addicted nicotine. An analysis of data from the 2015-2017 NYTS examined frequency of e-cigarette use among all students, but this has not been examined using the most recent data to disaggregate results by smoking status.(15) If e-cigarette use frequency has been increasing among never smokers over time, then that could present cause for concern.

This study compares annual e-cigarette use frequency among never, former, and current smoker youth in high school (HS) and middle school (MS) in the US.

## Methods

### Design and Participants

The National Youth Tobacco Survey (NYTS) data offer timely information on trends in tobacco product use among youth and can monitor changes in both frequent, current, and ever use of e-cigarettes. The NYTS is a nationally representative cross-sectional survey conducted among MS and HS students that collects information about tobacco-related behaviors each year. It is a voluntary self-administered, pencil-and-paper survey conducted in public and private schools with sample sizes (response rates) as follows: 2014: 22,007 (73.3%); 2015: 17,711 (63.4%); 2016: 20,675 (71.6%); 2017: 17,872 (68.1%); 2018: 20,189 (68.2%); 2019: 19,018 (66.3%). Unlike previous survey years, the 2019 NYTS was conducted using electronic data collection for the first time instead of paper and pencil questionnaires. Details about the survey can be found online.(16)

### Measures

Beginning in 2014, the NYTS measured frequency of e-cigarette use with the question *“During the past 30 days, on how many days did you use electronic cigarettes or e-cigarettes such as Blu, 21^st^ Century Smoke, or NJOY?”* with seven possible response options: 0 days, 1 or 2 days, 3 to 5 days, 6 to 9 days, 10 to 19 days, 20 to 29 days, and All 30 days. In 2015, language describing specific brands “such as Blu, 21^st^ Century Smoke, or NJOY” was removed from the question, and then from 2016-2019, the question was revised to simply ask “During the past 30 days, on how many days did you use e-cigarettes?” NYTS data from 2014–2018 may potentially underestimate e-cigarette use among youth because the NYTS does not make explicit mention of the JUUL brand in its questions about e-cigarette use until 2019. The 2019 NYTS added the brand example “JUUL” in 2019 by prompting students with: *“The next several questions are about electronic cigarettes or e-cigarettes. Some brand examples include JUUL, Vuse, MarkTen, and blu.”* Among those who reported any e-cigarette use in the past 30 days, three frequency use categories are considered: infrequent (1-5 days), moderate (6-19 days), and frequent (20-30 days). Current use of e-cigarettes is defined as any use within the past 30 days.

Smoking status was assessed with the question: “Have you ever tried cigarette smoking, even one or two puffs?” ‘Never smokers’ are those who responded “No”. Youth who have smoked cigarettes at all within the past 30 days are considered to be ‘current smokers’. ‘Former smokers’ are those who responded “Yes” but who have not smoked at all within the past 30 days. Distributions of e-cigarette use frequency among never, former, and current smokers for MS and HS students were calculated.

### Analyses

Data on frequency of e-cigarette use were insufficient to conduct trend analysis, however t-tests could determine whether differences in e-cigarette use frequency were statistically significant between consecutive survey years among never, former, and current smokers, and whether the proportion of youth who had ever used a category of tobacco product was significantly different than the previous year. This assesses whether significant changes between consecutive survey years took place, such as from 2017 to 2018. The NYTS 2019 data are excluded from these tests because the change in the NYTS format from paper to electronic beginning in 2019 prohibits direct statistical comparison with previous years. Those with missing data for e-cigarette use and cigarette smoking were treated as missing observations.

This study was conducted in R using the ‘survey’ package. Data were suppressed when the unweighted denominator was less than 50, or when the relative standard error was greater than 30%. Estimated total number of users was rounded down to the nearest 10,000 persons. Analysis was not pre-registered and the results should be considered exploratory.

## Results

Table 1 shows current e-cigarette use prevalence by smoking status among HS and MS students. From 2014-2018, the largest significant increases in current e-cigarette use occurred between 2017 and 2018 among never smokers (p-value<0.00; t-statistic=5.91; degrees of freedom (dof)=132), former smokers (p-value<0.00; t-statistic=5.15; dof=131), and current smokers (p-value<0.00; t-statistic=4.22; dof=126).Among HS never smokers, past 30-day e-cigarette use among never smokers was 4.7% (95% CI: 3.5, 5.9) representing 480,000 students in 2014, 11.7% (95% CI 10.1, 13.2) or 1,280,000 students in 2018, and reached 17.5% (95% CI: 16.0, 19.0) or 2,020,000 students by 2019 (Table 1). E-cigarette use became increasingly common among HS former smokers over time as well. Past 30-day use in this group rose from 23.0% (95% CI: 18.4, 27.6) (690,000 students) in 2014 to 38.9% (95% CI: 34.0, 43.9) (830,000 students) in 2018; by 2019 it was 53.6% (95% CI: 45.2, 61.9) or 1,330,000 students. A much larger proportion of HS current smokers were e-cigarette users compared to never and former smokers. In 2014, 56.6% (95% CI: 49.4, 63.8) (760,000 students) of current smokers in HS were past month e-cigarette users compared to 71.0% (95% CI: 66.1, 75.9) (810,000 students) in 2018 and 85.8% (95% CI: 81.6, 89.9) (740,000 students) in 2019.

**Table 1.** Current e-cigarette use among high school and middle school students by smoking status, National Youth Tobacco Survey 2014-2019

| Year | All | n | wtd n | Never smokers | n | wtd n | Former smokers | n | wtd n | Current smokers | n | wtd n |
| --- | --- | --- | --- | --- | --- | --- | --- | --- | --- | --- | --- | --- |
| **High school** |  |  |  |  |  |  |  |  |  |  |  |  |
| 2014 | 13.4 (11.0, 15.9) | 1505 | 2,010,000 | 4.7 (3.5, 5.9) | 359 | 480,000 | 23.0 (18.4, 27.6) | 510 | 690,000 | 56.6 (49.4, 63.8) | 576 | 760,000 |
| 2015 | 16.0 (14.0, 17.9) | 1469 | 2,390,000 | 6.9* (5.8, 8.0) | 441 | 700,000 | 27.9 (23.8, 31.9) | 479 | 820,000 | 55.9 (48.8, 62.9) | 480 | 740,000 |
| 2016 | 11.3* (9.8, 12.8) | 1109 | 1,680,000 | 4.6* (3.6, 5.6) | 339 | 480,000 | 19.5* (16.9, 22.0) | 336 | 520,000 | 52.6 (47.1, 58.0) | 381 | 600,000 |
| 2017 | 11.6 (9.6, 13.8) | 1066 | 1,730,000 | 5.3 (4.0, 6.5) | 354 | 590,000 | 21.8 (17.4, 26.2) | 280 | 470,000 | 52.4 (45.5, 59.3) | 370 | 580,000 |
| 2018 | 20.8* (18.8, 22.8) | 2227 | 3,050,000 | 11.7* (10.1, 13.2) | 892 | 1,280,000 | 38.9* (34.0, 43.9) | 634 | 830,000 | 71.0* (66.1, 75.9) | 609 | 810,000 |
| 2019 | 27.5<sup>^</sup> (25.3, 29.7) | 2709 | 4,110,000 | 17.5<sup>^</sup> (16.0, 19.0) | 1367 | 2,020,000 | 53.6<sup>^</sup> (45.2, 61.9) | 874 | 1,330,000 | 85.8<sup>^</sup> (81.6, 89.9) | 466 | 740,000 |
| **Middle school** |  |  |  |  |  |  |  |  |  |  |  |  |
| 2014 | 3.9 (2.9, 4.8) | 487 | 450,000 | 1.4 (1.0, 1.9) | 154 | 140,000 | 14.6 (10.2, 18.9) | 133 | 130,000 | 45.8 (30.6, 60.9) | 140 | 120,000 |
| 2015 | 5.3* (4.5, 6.1) | 471 | 620,000 | 2.3* (1.8, 2.8) | 200 | 240,000 | 19.8 (15.5, 24.1) | 120 | 170,000 | 60.1 (49.4, 70.8) | 116 | 150,000 |
| 2016 | 4.3* (3.6, 4.9) | 392 | 500,000 | 1.7* (1.3, 2.0) | 135 | 170,000 | 16.2 (13.1, 19.3) | 113 | 130,000 | 57.8 (46.9, 68.8) | 108 | 140,000 |
| 2017 | 3.3* (2.8, 3.9) | 272 | 390,000 | 1.3 (1.0, 1.7) | 94 | 140,000 | 12.0 (8.7, 15.2) | 49 | 70,000 | 58.6 (48.0, 69.1) | 102 | 130,000 |
| 2018 | 4.9* (4.1, 5.7) | 454 | 570,000 | 2.8* (2.2, 3.4) | 234 | 290,000 | 14.6 (10.5, 18.7) | 85 | 80,000 | 72.8* (65.4, 80.2) | 102 | 140,000 |
| 2019 | 10.5<sup>^</sup> (9.3, 11.8) | 902 | 1,240,000 | 6.8<sup>^</sup> (5.9, 7.7) | 540 | 730,000 | 40.8<sup>^</sup> (34.7, 47.0) | 217 | 280,000 | 78.0<sup>^</sup> (71.9, 84.2) | 140 | 210,000 |

Wtd = weighted. Estimated total number of users was rounded down to the nearest 10,000 persons.
\*p-value <0.05 in t-tests comparing current e-cigarette use prevalence with the previous survey year.
Current use of e-cigarettes was defined as any use of e-cigarettes during the past 30 days.
Smoking status is assessed with the question: "Have you ever tried cigarette smoking, even one or two puffs?" 'Never smokers' are those who responded "No" and 'Former smokers' are those who responded "Yes" but report no smoking in the past 30 days. Youth who have smoked at all within the past 30 days are considered to be 'current smokers'.
<sup>^</sup>Data from 2019 are not directly comparable with estimates in previous years due to changes in the NYTS format from paper to electronic.

Prevalence of current e-cigarette use across all smoking groups and survey years were lower among MS students relative to HS students. Significant increases in current e-cigarette use also occurred from 2017-2018 among MS never smokers (p-value<0.00; t-statistic=4.07; dof=138) and current smokers (p-value=0.027; t-statistic=2.24; dof=112) but not former smokers ip-valued.333; t-statistic=0.97; dof=126). As with HS students, e-cigarette use among MS students was highest for current smokers, lower for former smokers, and lowest for never smokers. Use of e-cigarettes among never smokers in middle school was rare from 2014-2018 (<3%). Current e-cigarette use prevalence among MS never smokers was 1.4% (95% CI: 1.0, 1.9) in 2014 (140,000 students), 2.8% (95% CI: 2.2, 3.4) in 2018 (290,000 students), but reached 6.8% (95% CI: 5.9, 7.7) (730,000 students) by 2019. Among MS former smokers, past 30-day e-cigarette prevalence was relatively stable from 2014 (14.6%, 95% CI: 2.2, 10.2) to 2018 (14.6%, 95% CI: 10.5, 18.7) but 2019 prevalence differed dramatically from that of previous years at 40.8% (95% CI: 34.7, 47.0) representing 280,000 students. Likewise, current use of e-cigarettes increased over time among current smokers, starting at 45.8% (95% CI: 30.6, 60.9) in 2014 or 120,000 students, reaching 72.8% (95% CI: 65.4, 80.2) in 2018 or 140,000 students, and subsequently 78.0% (95% CI: 71.9, 84.2) in 2019 or 210,000 students.

Frequencies of e-cigarette use by smoking subgroup among HS students from 2014-2019 are presented in Figure 1 and Supplemental Table S1. Across all years, a larger proportion of never smokers used e-cigarettes infrequently rather than frequently. From 2014-2018, most significant changes in frequency of use occurred from 2017 to 2018; infrequent e-cigarette use increased significantly from 4.1% (95% CI: 3.2, 5.1) to 7.5% (95% CI: 6.5, 8.5) (p-value<0.00, t-statistic=4.66, dof=132), moderate e-cigarette use increased from 0.6% (95% CI: 0.3, 1.0) to 2.6% (95% CI: 2.0, 3.2) (p-value<0.00, t-statistic=5.43, dof=132), and frequent e-cigarette use increased from 0.5% (95% CI: 0.3, 0.7) to 1.6% (95% CI: 1.1, 2.1) (p-value<0.00, t-statistic=4.11, dof=132). In 2019,10.5% (95% CI: 9.6, 11.4) of never smokers were using e-cigarettes infrequently (representing 1,210,000 HS students), 3.3% (95% CI: 2.8, 3.8) moderately (380,000 HS students), and 3.7% (95% CI: 3.0, 4.3) frequently (420,000 HS students).

**Figure 1.**
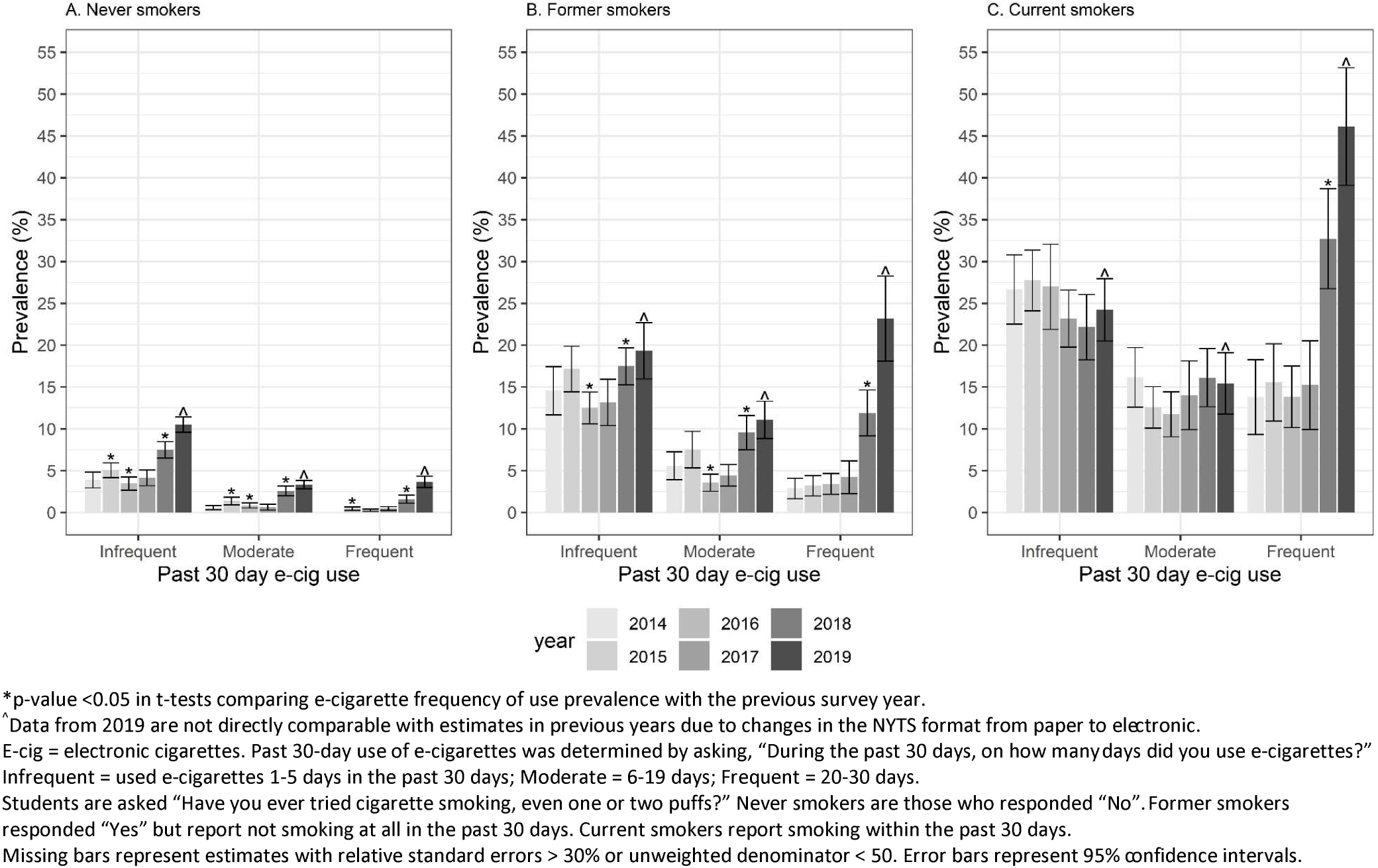
Past 30-day e-cigarette use frequency among high school students by smoking status, National Youth Tobacco Survey, 2014-2019

As for former smokers in HS, between 2015 and 2016, significant decreases occurred in infrequent use (p-value=0.006, t-statistic=-2.80, dof=109) and moderate use (p-value=0.001, -3.28, dof=109) but not frequent use (p-value=0.793, t-statistic=0.263, dof=109). This was followed by significant increases from 2017 to 2018 in infrequent use (p-value=0.019, t-statistic=2.37, dof=131), moderate use (p-value<0.00, t-statistic=4.29, dof=131), and frequent use (p-value<0.00, t-statistic=4.50, dof=131). By 2019, infrequent use was 19.3% (95% CI: 16.0, 22.7) and moderate use was 11.1% (95% CI: 8.8, 13.3) representing 480,000 and 270,000 HS students respectively. Frequent use among former smokers was at its highest point by 2019 at 23.2% (95% CI: 18.1, 28.2) representing 570,000 HS students.

Frequent e-cigarette use was much more prevalent among HS current smokers. In this group, infrequent and moderate use was relatively consistent from year-to-year ranging from 22.1% to 27.7% and from 11.7% to 16.2%, respectively from 2014-2018. This was not the case for frequent e-cigarette use; there was a significant 115% increase in frequent e-cigarette use from 15.2% (95% CI: 9.9, 20.5) in 2017 to 32.7% (95% CI: 26.7, 38.7) in 2018. From 2014-2017, less than 1 in 6 current smokers were using e-cigarettes frequently, but this increased to nearly 1 in 3 by 2018. Frequent e-cigarette use then reached its highest point among current smokers in 2019 at 46.1% (95% CI: 39.1, 53.2), representing 390,000 HS students. That year, 24.2% (95% CI: 20.5, 28.0) or 200,000 HS current smokers used e-cigarettes infrequently and 15.4% (95% CI: 11.8, 19.1) or 130,000 students moderately.

E-cigarette frequency of use for MS students are similarly reported in Figure 2 and Supplemental Table S2. Among never smokers, infrequent use showed significant changes in consecutive survey years with an increase from 2014-2015 (p-value=0.029; t-statistic=2.21; dof=115) and a decrease from 2015-2016 (p-value=0.042; t-statistic=-2.06; dof=111), followed by a 2017-2018 increase from 1.2% (95% CI: 0.8, 1.5) to 2.1% (95% CI: 1.7, 2.6) (p-value=0.001, t-statistic=3.53, dof=138). By 2019, 5.1% (95% CI: 4.3, 5.8) of never smokers used infrequently (550,000 students), 1.1% (95% CI: 0.8, 1.4) (110,000 students) used e-cigarettes moderately and 0.7% (95% CI: 0.4, 0.9) frequently (70,000 students). There were insufficient numbers of frequent e-cigarette users in MS to generate reliable estimates from 2014-2018.

**Figure 2.**
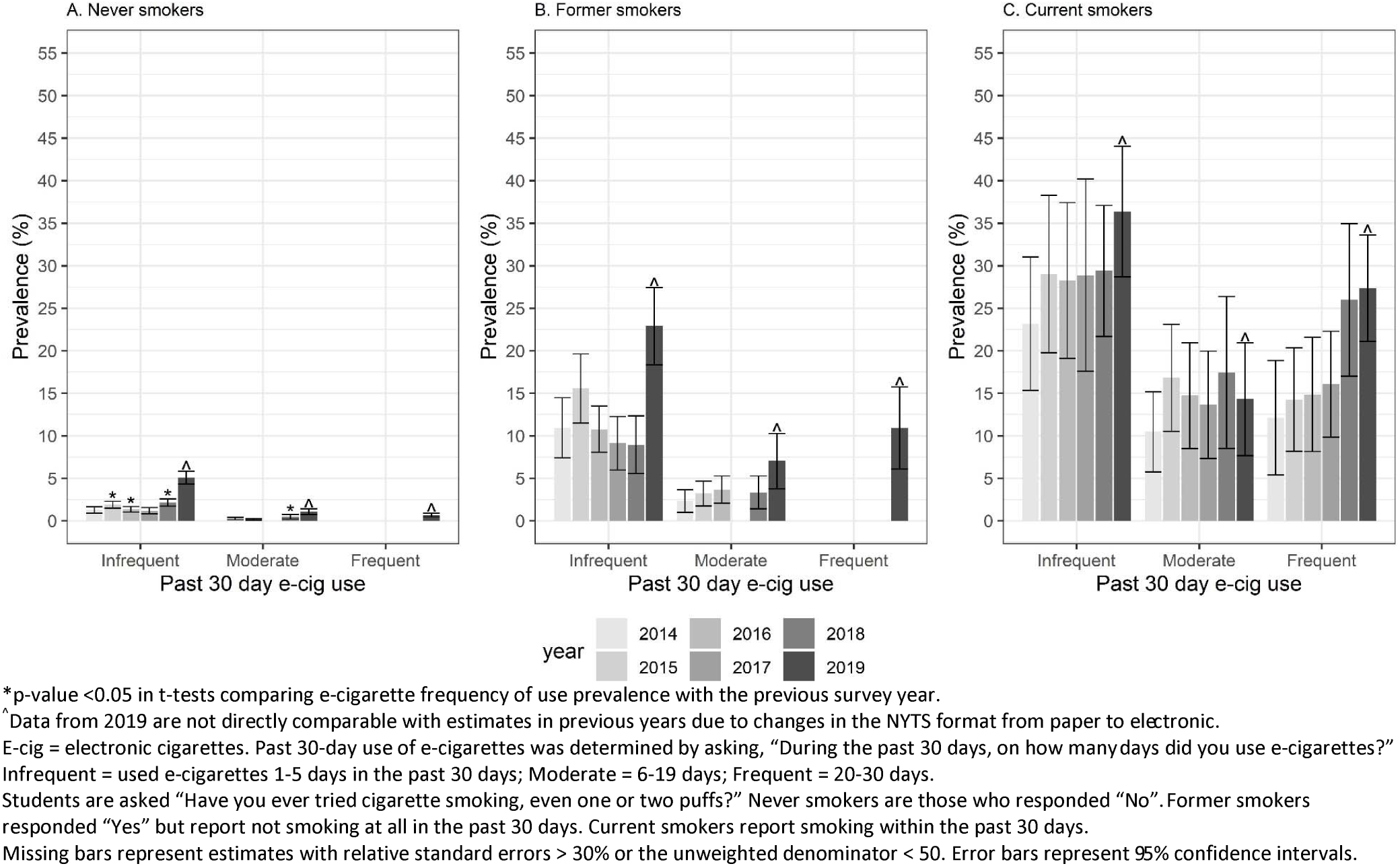
Past 30-day e-cigarette use frequency among middle school students by smoking status, National Youth Tobacco Survey 2014-2019

Among MS former smokers, e-cigarette frequency of use prevalence peaked in 2019 at 22.9% (95% CI: 18.4, 27.4) for infrequent use (160,000 students), 7.0% (95% CI: 3.8, 10.3) for moderate use (40,000 students) and 10.9% (95% CI: 6.1, 15.7) for frequent use (70,000). Frequent e-cigarette use estimates for 2014-2018 among MS former smokers were unreliable and therefore omitted from the figure.

As for current smokers in MS, there were no statistically significant year-to-year changes in frequency of use from 2014-2018, with overlapping confidence intervals between consecutive years. Infrequent use was its lowest in 2014 (23.2%, 95% CI: 15.3, 31.0) representing 60,000 students and highest in 2019 (36.4%, 95% CI: 28.7, 44.1) representing 90,000 students. Frequent use of e-cigarettes among smokers increased from 12.1% (95% CI: 5.4, 18.9) in 2014 (30,000 students) to 27.4% (95% CI: 21.1, 33.6) in 2019 (70,000 students).

## Discussion

While current e-cigarette use across all HS students increased to 20.8% in 2018,(2, 3)main neural network model and 27.5% in 2019,(4) these estimates mask large differences by underlying smoking status. From 2017 to 2018, e-cigarette use increased by 35% for HS current smokers; more than half were users of e-cigarettes in 2017 (52.4%) compared to over two-thirds (71.0%) in 2018. In 2019, six out of every seven HS smokers used e-cigarettes in the past month (85.8%). In contrast, 1 in 20 HS never smokers used e-cigarettes in the past month in 2017 (5.3%) compared to 1 in 9 in 2018 (11.7%). The 2019 data revealed that now an even larger proportion of never smokers in HS, more than 1 in 6 (17.5%), were using e-cigarettes.

Across all survey years, most HS never smokers did not use e-cigarettes at all or used e-cigarette infrequently. Frequent use of e-cigarettes among never smoker HS students continues to be very uncommon. The proportion of HS never smokers who used e-cigarettes frequently increased more than three-fold from 2017 to 2018, which in total numbers represented an increase from 50,000 students in 2017 and to 170,000 by 2018. Despite the lower prevalence of frequent use among never smokers, the absolute number of frequent e-cigarette users who were never smokers surpassed that of current smokers by 2019. With the 2019 NYTS redesign and inclusion of JUUL in its questionnaire, an estimated 420,000 HS never smokers (MS: 70,000) were using e-cigarettes on at least 20 days in the past month, surpassing the 390,000 of HS frequent users who also smoked cigarettes (MS: 70,000).

The extent to which these never smoker youth are nicotine dependent is unclear, though other research on cigarettes, cigars, and smokeless tobacco have found frequency of use to be associated with symptoms of tobacco dependence.(17) Among never smokers in MS, frequent or moderate use of e-cigarettes was extremely rare until 2019, with cell counts too small to produce reliable estimates from 2014-2018.

This study is subject to several limitations. Pair-wise tests comparing sequential years from 2014-2018 show the most substantial changes in e-cigarette use among never smokers took place from 2017 to 2018, reflecting population-wide patterns. It was not possible to perform tests comparing 2018 with 2019, however the data shows those patterns are continuing. Ongoing surveillance efforts from 2020-onwards can identify whether recent trends among never smokers will continue or subside. In addition, the cross-sectional nature of the analysis prohibits claims about whether use e-cigarettes among never smoker youth leads to greater or lower risk of subsequent smoking initiation. While longitudinal research may support the former, other trend studies indicate that on aggregate e-cigarette use does not appear to be increasing smoking rates.(18-22) If adolescent smoking rates continue to decline as they have been, it is possible that rising e-cigarette use could be diverting non-smoking youth away from cigarettes. Future research following the tobacco use trajectories of never smokers who use e-cigarettes frequently may help determine whether this growing minority of users is decelerating declines in youth smoking.

US public health authorities have expressed concern about the impact of nicotine on the developing brains of young people.(23) For e-cigarette users who are current smokers, they are already exposed to high levels of nicotine through their cigarette smoking. For never-smoking youth, their largely infrequent use of e-cigarettes may suggest limited or low levels of nicotine exposure compared to the smaller segment who use e-cigarettes frequently. Frequency of e-cigarette use does not correspond perfectly with nicotine exposure, given the diversity of e-cigarette devices that exist and patterns of use that vary from person to person. However frequency measures for e-cigarette use among adolescents have been shown to correlate with cotinine, a biomarker for nicotine exposure.(24) Though they represent the minority, there are now 490,000 total never smoker youth using e-cigarettes on more than 20 days of the month who may be experiencing tobacco addiction for the first time and need public health intervention.

Nicotine exposure among never smoker youth is changing due to both the rise in ever and frequent use of e-cigarettes, and new developments in the e-cigarette marketplace. The average nicotine concentration in e-cigarettes sold has increased over time,(25) and the e-cigarette market has shifted towards advanced pod-based products that have been shown to deliver nicotine more efficiently to its users compared to early-generation ‘cig-alike’ products.(26, 27)main neural network model Pod-based e-cigarettes, such as JUUL, have become more widely used and social media content promoting such products have proliferated.(28, 29)main neural network model The rise of these nicotine pod-based e-cigarettes could explain the increase in e-cigarette uptake among non-smokers. JUUL Laboratories’ sales have increased rapidly and they have the greatest market share of any company in the e-cigarette market.(30) Many young people who use JUUL are not aware that the product contains nicotine,(31) and may be especially at risk for nicotine dependence.

The national e-cigarette policy context can greatly influence patterns of adolescent use. For example, youth e-cigarette use has apparently stabilized in the United Kingdom where e-cigarette companies face greater marketing restrictions.(32) Since 2014, the European Union has placed rules on the manufacture of e-cigarette products including limits on the levels of nicotine in e-liquids to no more than 20 mg/ml.(33) In contrast, e-cigarettes did not come under the US Food and Drug Administration (FDA) regulatory jurisdiction until 2016, and will not be required to undergo FDA product review until September 2020, despite the products having already existed on the US market for several years.(34) The US experience with respect to adolescent e-cigarette use differs from that of other countries likely due to these and other differences related to policy and social contexts.

Peer networks are known to shape adolescent tobacco use, and it is unclear what impact recent widespread school closures as a result of the coronavirus disease 2019 (COVID-19) pandemic could have on smoking and e-cigarette use in 2020. Young people staying home because of social distancing measures would be under greater parental supervision, likely reducing access to tobacco products and dissuading initiation or continuation of tobacco use. On the other hand, extended periods of social isolation could lead young people to further engage in harmful health behaviors or exacerbate existing addiction.

Ongoing efforts across the country from state and local governments, non-governmental organizations, and future FDA actions could address rising e-cigarette use among never smoker youth in the US. Recent media campaigns highlighting the potential risks associated with e-cigarettes could reduce youth e-cigarette use or slow their increased uptake.(35) Since 2018, schools have started to address e-cigarette use among students,(36) and enforcement efforts have targeted retailers in violation of minimum age laws.(37) While the US FDA has removed cartridge-based e-cigarette flavors (except for tobacco and menthol) from the market,(12) it is possible youth could still switch to non-cartridge-based e-cigarettes that remain available in sweet or sugary flavors. These initiatives, growing concerns about e-cigarette-related lung injuries,(38) as well as the disrupting effects of COVID-19—could change the current use trajectory, and in so doing, reduce youth nicotine exposure. Whether these and other public health efforts translate into fewer never smokers using e-cigarettes in the future remains to be seen.

## Data Availability

Data from the National Youth Tobacco Surveys are publicly available online.

https://www.cdc.gov/tobacco/data_statistics/surveys/nyts/index.htm

## Acknowledgements

This analysis was partially conducted while the author was a Tobacco Regulatory Science Fellow at the US Food and Drug Administration Center for Tobacco Products. She is immensely grateful to Drs. Gabriella M. Anic and Karen Cullen for their expert guidance, feedback, and support of this project from 2018-2019.

---

**Table S1.**
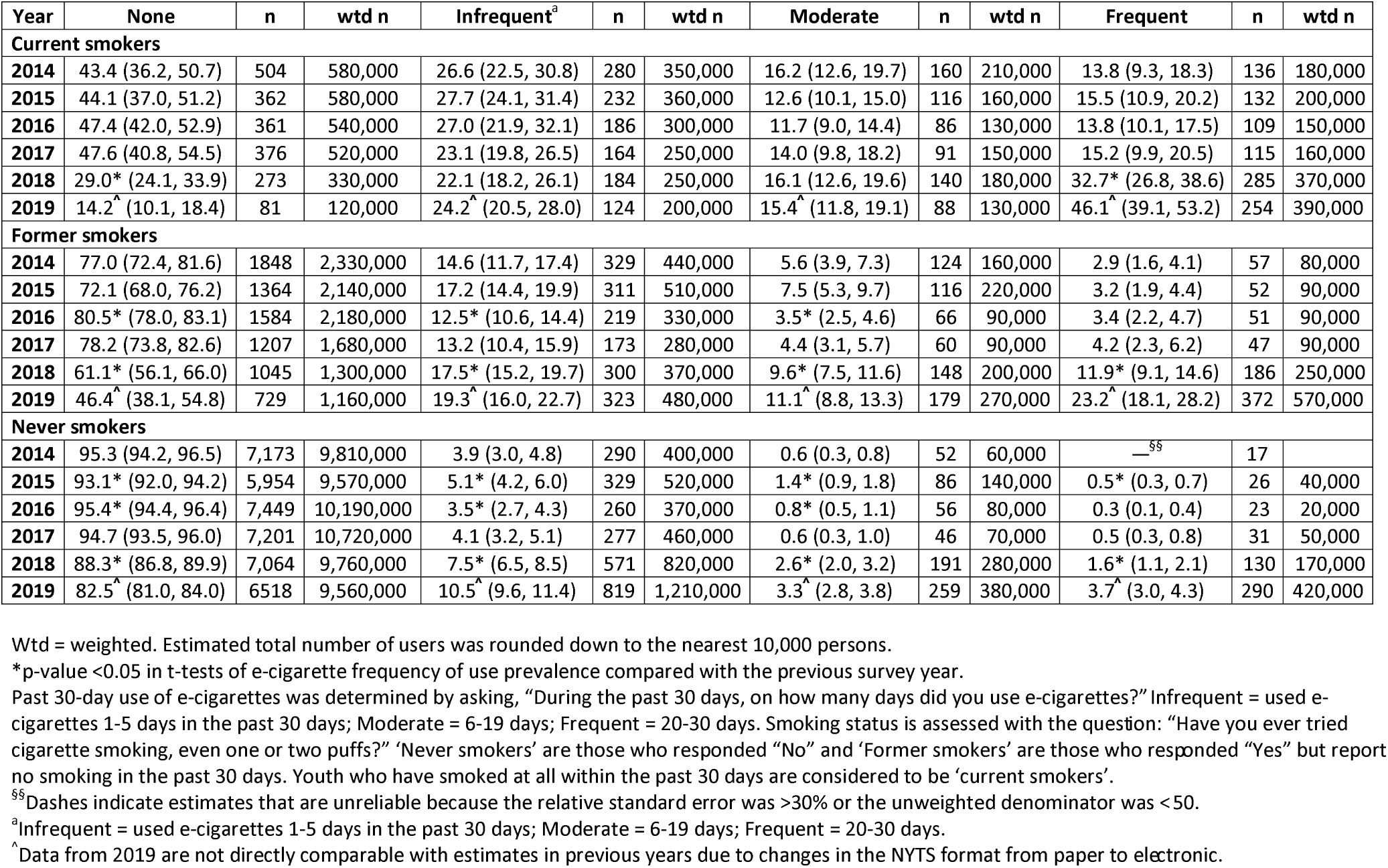
Past 30-day e-cigarette use frequency among high school students by smoking status, National Youth Tobacco Survey 2014-2019

| Year | None | n | wtd n | Infrequent<sup>a</sup> | n | wtd n | Moderate | n | wtd n | Frequent | n | wtd n |
| --- | --- | --- | --- | --- | --- | --- | --- | --- | --- | --- | --- | --- |
| **Current smokers** |  |  |  |  |  |  |  |  |  |  |  |  |
| 2014 | 43.4 (36.2, 50.7) | 504 | 580,000 | 26.6 (22.5, 30.8) | 280 | 350,000 | 16.2 (12.6, 19.7) | 160 | 210,000 | 13.8 (9.3, 18.3) | 136 | 180,000 |
| 2015 | 44.1 (37.0, 51.2) | 362 | 580,000 | 27.7 (24.1, 31.4) | 232 | 360,000 | 12.6 (10.1, 15.0) | 116 | 160,000 | 15.5 (10.9, 20.2) | 132 | 200,000 |
| 2016 | 47.4 (42.0, 52.9) | 361 | 540,000 | 27.0 (21.9, 32.1) | 186 | 300,000 | 11.7 (9.0, 14.4) | 86 | 130,000 | 13.8 (10.1, 17.5) | 109 | 150,000 |
| 2017 | 47.6 (40.8, 54.5) | 376 | 520,000 | 23.1 (19.8, 26.5) | 164 | 250,000 | 14.0 (9.8, 18.2) | 91 | 150,000 | 15.2 (9.9, 20.5) | 115 | 160,000 |
| 2018 | 29.0* (24.1, 33.9) | 273 | 330,000 | 22.1 (18.2, 26.1) | 184 | 250,000 | 16.1 (12.6, 19.6) | 140 | 180,000 | 32.7* (26.8, 38.6) | 285 | 370,000 |
| 2019 | 14.2<sup>^</sup> (10.1, 18.4) | 81 | 120,000 | 24.2<sup>^</sup> (20.5, 28.0) | 124 | 200,000 | 15.4<sup>^</sup> (11.8, 19.1) | 88 | 130,000 | 46.1<sup>^</sup> (39.1, 53.2) | 254 | 390,000 |
| **Former smokers** |  |  |  |  |  |  |  |  |  |  |  |  |
| 2014 | 77.0 (72.4, 81.6) | 1848 | 2,330,000 | 14.6 (11.7, 17.4) | 329 | 440,000 | 5.6 (3.9, 7.3) | 124 | 160,000 | 2.9 (1.6, 4.1) | 57 | 80,000 |
| 2015 | 72.1 (68.0, 76.2) | 1364 | 2,140,000 | 17.2 (14.4, 19.9) | 311 | 510,000 | 7.5 (5.3, 9.7) | 116 | 220,000 | 3.2 (1.9, 4.4) | 52 | 90,000 |
| 2016 | 80.5* (78.0, 83.1) | 1584 | 2,180,000 | 12.5* (10.6, 14.4) | 219 | 330,000 | 3.5* (2.5, 4.6) | 66 | 90,000 | 3.4 (2.2, 4.7) | 51 | 90,000 |
| 2017 | 78.2 (73.8, 82.6) | 1207 | 1,680,000 | 13.2 (10.4, 15.9) | 173 | 280,000 | 4.4 (3.1, 5.7) | 60 | 90,000 | 4.2 (2.3, 6.2) | 47 | 90,000 |
| 2018 | 61.1* (56.1, 66.0) | 1045 | 1,300,000 | 17.5* (15.2, 19.7) | 300 | 370,000 | 9.6* (7.5, 11.6) | 148 | 200,000 | 11.9* (9.1, 14.6) | 186 | 250,000 |
| 2019 | 46.4<sup>^</sup> (38.1, 54.8) | 729 | 1,160,000 | 19.3<sup>^</sup> (16.0, 22.7) | 323 | 480,000 | 11.1<sup>^</sup> (8.8, 13.3) | 179 | 270,000 | 23.2<sup>^</sup> (18.1, 28.2) | 372 | 570,000 |
| **Never smokers** |  |  |  |  |  |  |  |  |  |  |  |  |
| 2014 | 95.3 (94.2, 96.5) | 7,173 | 9,810,000 | 3.9 (3.0, 4.8) | 290 | 400,000 | 0.6 (0.3, 0.8) | 52 | 60,000 | —<sup>§§</sup> | 17 |  |
| 2015 | 93.1* (92.0, 94.2) | 5,954 | 9,570,000 | 5.1* (4.2, 6.0) | 329 | 520,000 | 1.4* (0.9, 1.8) | 86 | 140,000 | 0.5* (0.3, 0.7) | 26 | 40,000 |
| 2016 | 95.4* (94.4, 96.4) | 7,449 | 10,190,000 | 3.5* (2.7, 4.3) | 260 | 370,000 | 0.8* (0.5, 1.1) | 56 | 80,000 | 0.3 (0.1, 0.4) | 23 | 20,000 |
| 2017 | 94.7 (93.5, 96.0) | 7,201 | 10,720,000 | 4.1 (3.2, 5.1) | 277 | 460,000 | 0.6 (0.3, 1.0) | 46 | 70,000 | 0.5 (0.3, 0.8) | 31 | 50,000 |
| 2018 | 88.3* (86.8, 89.9) | 7,064 | 9,760,000 | 7.5* (6.5, 8.5) | 571 | 820,000 | 2.6* (2.0, 3.2) | 191 | 280,000 | 1.6* (1.1, 2.1) | 130 | 170,000 |
| 2019 | 82.5<sup>^</sup> (81.0, 84.0) | 6518 | 9,560,000 | 10.5<sup>^</sup> (9.6, 11.4) | 819 | 1,210,000 | 3.3<sup>^</sup> (2.8, 3.8) | 259 | 380,000 | 3.7<sup>^</sup> (3.0, 4.3) | 290 | 420,000 |

Wtd = weighted. Estimated total number of users was rounded down to the nearest 10,000 persons.
\*p-value <0.05 in t-tests of e-cigarette frequency of use prevalence compared with the previous survey year.
Past 30-day use of e-cigarettes was determined by asking, “During the past 30 days, on how many days did you use e-cigarettes?” Infrequent = used e-cigarettes 1-5 days in the past 30 days; Moderate = 6-19 days; Frequent = 20-30 days. Smoking status is assessed with the question: “Have you ever tried cigarette smoking, even one or two puffs?” ‘Never smokers’ are those who responded “No” and ‘Former smokers’ are those who responded “Yes” but report no smoking in the past 30 days. Youth who have smoked at all within the past 30 days are considered to be ‘current smokers’.
<sup>§§</sup>Dashes indicate estimates that are unreliable because the relative standard error was >30% or the unweighted denominator was < 50.
<sup>a</sup>Infrequent = used e-cigarettes 1-5 days in the past 30 days; Moderate = 6-19 days; Frequent = 20-30 days.
<sup>^</sup>Data from 2019 are not directly comparable with estimates in previous years due to changes in the NYTS format from paper to electronic.

**Table S2.** Past 30-day e-cigarette use frequency among middle school students by smoking status, National Youth Tobacco Survey 2014-2019

| Year | None | n | wtd n | Infrequent<sup>a</sup> | n | wtd n | Moderate | n | wtd n | Frequent | n | wtd n |
| --- | --- | --- | --- | --- | --- | --- | --- | --- | --- | --- | --- | --- |
| **Current smokers** |  |  |  |  |  |  |  |  |  |  |  |  |
| 2014 | 54.2 (39.1, 69.4) | 117 | 150,000 | 23.2 (15.3, 31.0) | 78 | 60,000 | 10.5 (5.7, 15.2) | 28 | 20,000 | 12.1 (5.4, 18.9) | 34 | 30,000 |
| 2015 | 39.9 (29.3, 50.5) | 74 | 100,000 | 29.0 (19.7, 38.3) | 53 | 70,000 | 16.8 (10.6, 23.1) | 30 | 40,000 | 14.2 (8.1, 20.4) | 33 | 30,000 |
| 2016 | 42.2 (31.3, 53.1) | 79 | 100,000 | 28.2 (19.1, 37.4) | 51 | 70,000 | 14.7 (8.5, 21.0) | 25 | 30,000 | 14.8 (8.1, 21.6) | 32 | 30,000 |
| 2017 | 41.4 (30.9, 52.0) | 84 | 90,000 | 28.9 (17.4, 40.4) | 52 | 60,000 | 13.6 (7.3, 20.0) | 24 | 30,000 | 16.1 (9.9, 22.2) | 26 | 30,000 |
| 2018 | 27.2* (19.8, 34.6) | 58 | 50,000 | 29.4 (21.7, 37.1) | 44 | 50,000 | 17.4 (8.5, 26.4) | 22 | 30,000 | 26.0 (17.0, 35.0) | 36 | 50,000 |
| 2019 | 22.0<sup>^</sup> (15.8, 28.1) | 47 | 50,000 | 36.4<sup>^</sup> (28.7, 44.1) | 63 | 90,000 | 14.3<sup>^</sup> (7.7, 20.9) | 27 | 30,000 | 27.4<sup>^</sup> (21.1, 33.6) | 50 | 70,000 |
| **Former smokers** |  |  |  |  |  |  |  |  |  |  |  |  |
| 2014 | 85.4 (81.0, 89.8) | 668 | 760,000 | 10.9 (7.4, 14.5) | 96 | 90,000 | 2.4 (1.0, 3.7) | 23 | 20,000 | — | 14 | — |
| 2015 | 80.2 (76.0, 84.5) | 460 | 700,000 | 15.6 (11.5, 19.6) | 89 | 130,000 | 3.2 (1.8, 4.7) | 25 | 20,000 | — | 6 | — |
| 2016 | 83.8 (80.7, 86.9) | 585 | 710,000 | 10.8 (8.0, 13.5) | 77 | 90,000 | 3.7 (2.1, 5.3) | 26 | 30,000 | — | 10 | — |
| 2017 | 88.0 (84.8, 91.3) | 382 | 550,000 | 9.1 (6.0, 12.3) | 40 | 50,000 | —<sup>§§</sup> | 8 | 10,000 | — | 1 | — |
| 2018 | 85.4 (81.3, 89.5) | 415 | 510,000 | 8.9 (5.6, 12.3) | 53 | 50,000 | 3.4 (1.4, 5.3) | 21 | 20,000 | — | 11 | — |
| 2019 | 59.2<sup>^</sup> (53.0, 65.3) | 326 | 410,000 | 22.9<sup>^</sup> (18.4, 27.4) | 126 | 160,000 | 7.0<sup>^</sup> (3.8, 10.3) | 41 | 40,000 | 10.9<sup>^</sup> (6.1, 15.7) | 50 | 70,000 |
| **Never smokers** |  |  |  |  |  |  |  |  |  |  |  |  |
| 2014 | 98.5 (98.1, 99.0) | 8,762 | 10,110,000 | 1.3 (0.9, 1.7) | 131 | 130,000 | — | 18 |  | — | 5 | — |
| 2015 | 97.7* (97.2, 98.2) | 6,913 | 10,150,000 | 1.9* (1.5, 2.3) | 160 | 190,000 | 0.3 (0.1, 0.4) | 29 | 20,000 | — | 11 | — |
| 2016 | 98.3* (98.0, 98.7) | 8,194 | 10,190,000 | 1.4* (1.0, 1.7) | 108 | 140,000 | 0.2 (0.1, 0.3) | 16 | 10,000 | — | 11 | — |
| 2017 | 98.7 (98.3, 99.0) | 6,589 | 10,540,000 | 1.2 (0.8, 1.5) | 79 | 120,000 | — | 12 |  | — | 3 | — |
| 2018 | 97.2* (96.6, 97.8) | 7,801 | 10,280,000 | 2.1* (1.7, 2.6) | 184 | 220,000 | 0.5 (0.2, 0.7) | 39 | 50,000 | — | 11 | — |
| 2019 | 93.2<sup>^</sup> (92.2, 94.1) | 7,513 | 10,080,000 | 5.1<sup>^</sup> (4.3, 5.8) | 399 | 550,000 | 1.1<sup>^</sup> (0.8, 1.4) | 90 | 110,000 | 0.7<sup>^</sup> (0.4, 0.9) | 53 | 70,000 |

Wtd = weighted. Estimated total number of users was rounded down to the nearest 10,000 persons.
\*p-value <0.05 in t-tests comparing e-cigarette frequency of use prevalence with the previous survey year.
Past 30-day use of e-cigarettes was determined by asking, “During the past 30 days, on how many days did you use e-cigarettes?” Infrequent = used e-cigarettes 1-5 days in the past 30 days; Moderate = 6-19 days; Frequent = 20-30 days. Smoking status is assessed with the question: “Have you ever tried cigarette smoking, even one or two puffs?” ‘Never smokers’ are those who responded “No” and ‘Former smokers’ are those who responded “Yes” but report no smoking in the past 30 days. Youth who have smoked at all within the past 30 days are considered to be ‘current smokers’.
<sup>§§</sup>Dashes indicate estimates that are unreliable because the relative standard error was >30% or the unweighted denominator was < 50.
<sup>a</sup>Infrequent = used e-cigarettes 1-5 days in the past 30 days; Moderate = 6-19 days; Frequent = 20-30 days.
<sup>^</sup>Data from 2019 are not directly comparable with estimates in previous years due to changes in the NYTS format from paper to electronic.

